# Telephone advice lines available out-of-hours to people with palliative and end-of-life care needs: a qualitative interview study with professionals and development of a practical framework to improve services

**DOI:** 10.1101/2023.10.19.23297190

**Authors:** Sophie Pask, Allen Omoruyi, Ahmed Mohamed, Rachel L. Chambers, Phillippa G. McFarlane, Therese Johansson, Rashmi Kumar, Andy Woodhead, Ikumi Okamoto, Stephen Barclay, Irene J. Higginson, Katherine E. Sleeman, Fliss E. M. Murtagh

## Abstract

**Background:** People living at home with advanced illness require around-the-clock care. Telephone-based advice lines are critical for accessing help, yet evidence is limited.

**Aim:** To explore ‘out-of-hours’ telephone-based advice lines available to adults living at home with advanced illness and their carers across the UK, and construct a practical framework to improve services.

**Design:** Structured qualitative interviews with thematic analysis. A patient and public involvement workshop was conducted to refine the proposed framework.

**Setting/participants:** Professionals with palliative/end-of-life care commissioning responsibilities, or knowledge of out-of-hours service provision, were purposively sampled to ensure UK-wide representation.

**Results:** Seventy-one interviews were conducted, covering 60 geographical areas. Five themes were identified. *Availability:* Ten models of advice lines were described. Variation led to confusion about who to call and when. *Accessibility, awareness and promotion:* It was assumed that patients/carers know who to call out-of-hours, but often they did not. *Practicalities:* Call handlers skills/expertise varied, which influenced how calls were managed. Possible responses ranged from simply signposting to organising home visits. *Integration/continuity of care:* Integration between care providers was limited by electronic medical records access and information sharing. *Service structure/commissioning:* Sustained funding was often an issue for charitably funded organisations.

**Conclusions:** Multiple advice lines lead to confusion and delays in obtaining care, as many default to general ‘out-of-hours’ advice lines. Dedicated advice lines are valuable for patients with advanced illness as long as they are implemented well. A practical framework (including a comprehensive overview of components) is provided for guiding how these are delivered.

**Key statements:** What is already known about the topic?

- *People living at home with advanced illness and those that care for them need access to dedicated palliative and end-of-life care 24 hours a day, 7-days a week*.
- *While understanding of telephone advice lines often exists at a single service level, there is limited knowledge in terms of national provision*.

What does this paper add?

- *This qualitative study provides an understanding of multiple telephone-based advice line services available out-of-hours at a national-level, and identifies a lack of consistency and challenges with integration between available services*.
- *Promotion of dedicated advice lines (or an area equivalent) needs to ensure that people with advanced illness are aware of how to access such support, but there is variation in how this is done*.
- *Incorporating the views of patients with advanced illness and carers in the development of telephone-based advice is essential to ensure the care delivered is centred around their needs*.

Implications for practice, theory or policy

- *The practical framework developed in this research (using key considerations from professionals based on structured interviews and a patient and public involvement workshop) can be used to guide commissioners and service providers*.

## Background

Ageing populations, and the rapid projected rise in multiple health conditions, present global challenges for palliative care.^1^ Demand for community-based palliative care is anticipated to increase worldwide,^1^ but resources are lacking.^2^ Palliative care has demonstrable benefits for people with advanced illness,^3–5^ and to healthcare systems (where modest investments in community-based care have enabled fewer deaths in hospital).^6–10^

Community-based palliative care is provided by numerous professionals with variable training and expertise.^3, 11, 12^ This includes primary and community care teams (e.g. GPs and district/community nurses), social care teams (e.g. home care workers), and specialist palliative care teams.^13^ Increasing community-based provision is necessary for the growing demand and to reduce pressure on acute services,^1, 14^ but is challenging and variable.^2, 5, 15, 16^

Out-of-hours services (i.e. overnight, and weekends or public holidays) are responsible for providing healthcare for two-thirds of the week.^17, 18^ Unscheduled out-of-hours care (e.g. symptom management) is often needed,^19, 20^ and has been identified as a priority for patients, carers and policymakers.^21^ One means of providing round-the-clock palliative care is via dedicated telephone advice lines, which aim to provide immediate access to adequate support.^14, 22^

People with advanced illness and their carers should have access to dedicated telephone-based advice and support 24/7.^23^ In the UK, this has been recommended as a minimum requirement by the National Institute for Health and Care Excellence (NICE) since 2004.^24^ Nearly two decades later, dedicated advice lines are still not universally available despite being a priority to achieve.^2, 25, 26^ Our Better End of Life research report found that almost a third of areas surveyed had no access to dedicated advice lines out-of-hours, and 42% reported challenges or limitations.^2^

## Aim

To explore ‘out-of-hours’ telephone-based advice lines available to adults living at home with advanced illness and their carers across the UK, and construct a practical framework to improve services.

### Objectives

(1) To explore professional perspectives on provision and delivery of out-of-hours telephone advice lines available for adults with advanced illness, and their carers, including gaps and variations in care.
(2) To construct a practical framework of key considerations for improving telephone advice line services.

## Methods

We report this study according to the Standards for Reporting Qualitative Research (Supplementary file 1),^27^ and patient and public involvement (PPI) in line with the GRIPP2 – Short Form^28^ (Supplementary file 2).

**Box 1. Defining telephone advice lines**

What do we mean by an ‘advice line’?

A number of terms have been used to describe telephone advice and support, such as ‘advice line’, ‘support line’ or ‘helpline’.

In this paper, ‘advice line’ will be used to refer to advice and support that adults living at home with advanced illness, and their carers, can access via telephone.

This encompasses telephone advice lines that are specifically for palliative and end-of-life care needs (otherwise referred to as a *<u>dedicated advice line</u>*), or more general advice lines (such as emergency services, non-emergency medical lines or primary care lines accessed out-of-hours). Advice lines may therefore be provided by non-specialists/administrators, primary care and community nursing services or specialist services.

The types of responses and actions that may be provided include: signposting (i.e. providing the contact details of other services that can help), triaging (i.e. process by which services inform the order and priority of care, or direct to the appropriate person), advice, emotional and psychological support, home visits, follow-up or referrals.

### Study design

Structured qualitative interviews as part of a cross-sectional survey across the UK. The full survey data is summarised elsewhere (see [Better End of life Report](https://www.mariecurie.org.uk/globalassets/media/documents/policy/beol-reports-2022/better-end-of-life-report-2022.pdf)).^2^

### Sampling and recruitment

Professionals with palliative and end-of-life care commissioning responsibilities, or good knowledge of out-of-hours service provision in the area were purposively sampled to ensure representation across each nation, and accompanied by a snowball approach.

Representation was sought across 42 Integrated Care Systems (England), seven Health Boards (Wales), 14 regional Health Boards (Scotland), and five Health and Social Care Trusts (Northern Ireland).

### Data collection

The research team contacted palliative and end-of-life care networks in each nation to share an email invitation to take part in the study, alongside a participant information sheet.

Interviews were scheduled at a suitable date/time via telephone or video link with those wishing to take part. Verbal consent was obtained using a consent script prior to conducting the interview. Interviews were audio recorded using an encrypted digital voice recorder. A topic guide (see Appendix 1) was developed around aspects of out-of-hours care that have been identified as important by patients, carers and professionals.^29^ Data were collected between 21st December 2021 and 1st June 2022. Interviews were conducted by a non-clinical postgraduate research associate experienced in qualitative research (SP), and previously known in a professional capacity to two participants recruited. SP transcribed several interviews (alongside AM, RLC, and PGM) or reviewed for clarity, which ensured immersion in the data. Anonymised electronic copies of transcripts were sent to participants for respondent checking.

### Data analysis

Interviews were analysed by SP, with support from AO and FEMM. NVivo^30^ was used to manage data. Interviews were analysed using Braun and Clarke’s^31^ method of thematic analysis, including: (1) data familiarisation, (2) generation of codes, (3) generation of initial themes, (4) developing and reviewing themes, (5) refining, defining and naming themes, and (6) the written report.^31^ An iterative approach was used rather than treating phases as distinct and unidirectional.^31^ Candidate themes were discussed with the wider research team, as well as during the refining, defining and naming stage.

### Patient and public involvement

As this research presented the views of professionals, we worked with a PPI group to ensure insights from people living with advanced illness and carers were included. A workshop was held online with thirteen patients and/or carers to review findings, and bring the lived experience perspective to help shape these, using an independent facilitator in June 2023 (see Supplementary File 3 for more details).

### Ethical approval

Ethical approval was granted by the Hull York Medical School Ethics Committee (Reference 21/22 7).

## Results

Interviews were conducted with 71 professionals, relating to 60 geographical areas across the UK (see Appendix 2). Five themes were identified.

### Availability of advice lines

#### Variation in the types of advice lines available

Professionals described ten distinct types of out-of-hours advice lines (see Table 2). The route to accessing help out-of-hours was complicated by the variety of different advice lines available, which often resulted in defaulting to national provision that addressed all urgent health issues (e.g. NHS 111/999).

**Table 1.**
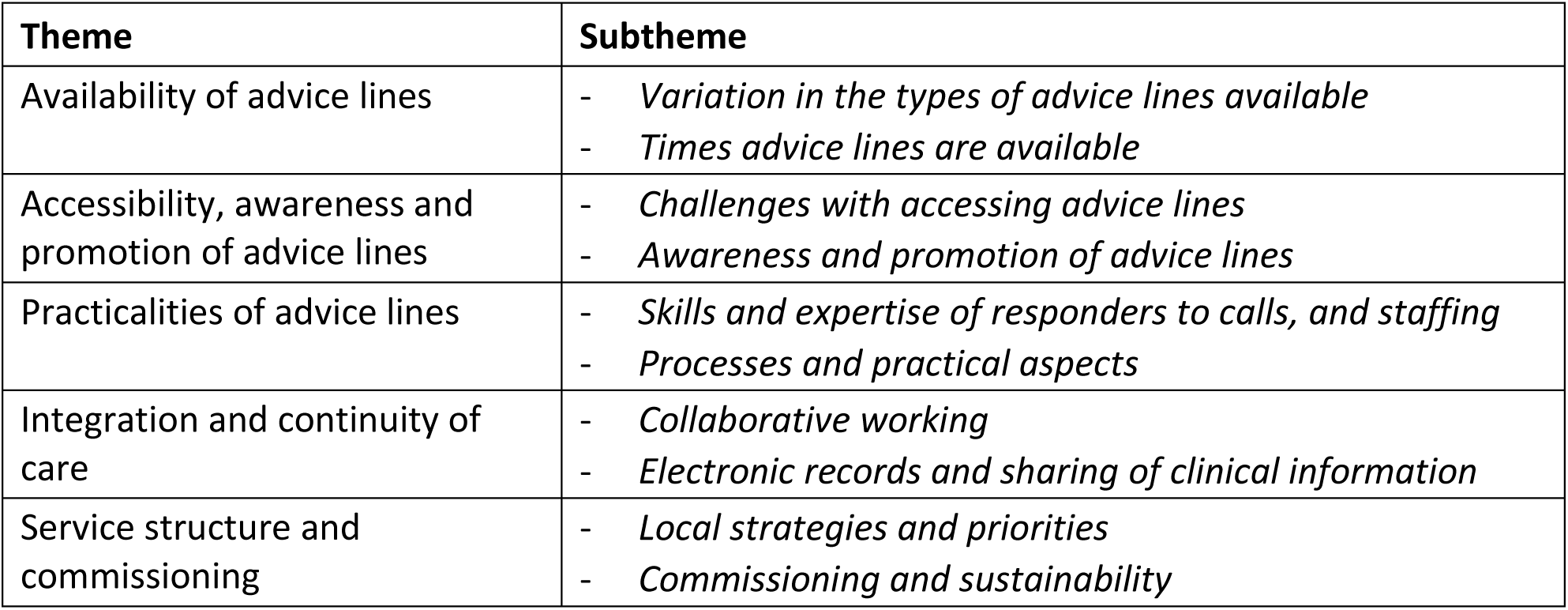
Summary of themes and subthemes.

| **Theme** | **Subtheme** |
| --- | --- |
| Availability of advice lines | - *Variation in the types of advice lines available*<br>- *Times advice lines are available* |
| Accessibility, awareness and promotion of advice lines | - *Challenges with accessing advice lines*<br>- *Awareness and promotion of advice lines* |
| Practicalities of advice lines | - *Skills and expertise of responders to calls, and staffing*<br>- *Processes and practical aspects* |
| Integration and continuity of care | - *Collaborative working*<br>- *Electronic records and sharing of clinical information* |
| Service structure and commissioning | - *Local strategies and priorities*<br>- *Commissioning and sustainability* |

**Table 2.**
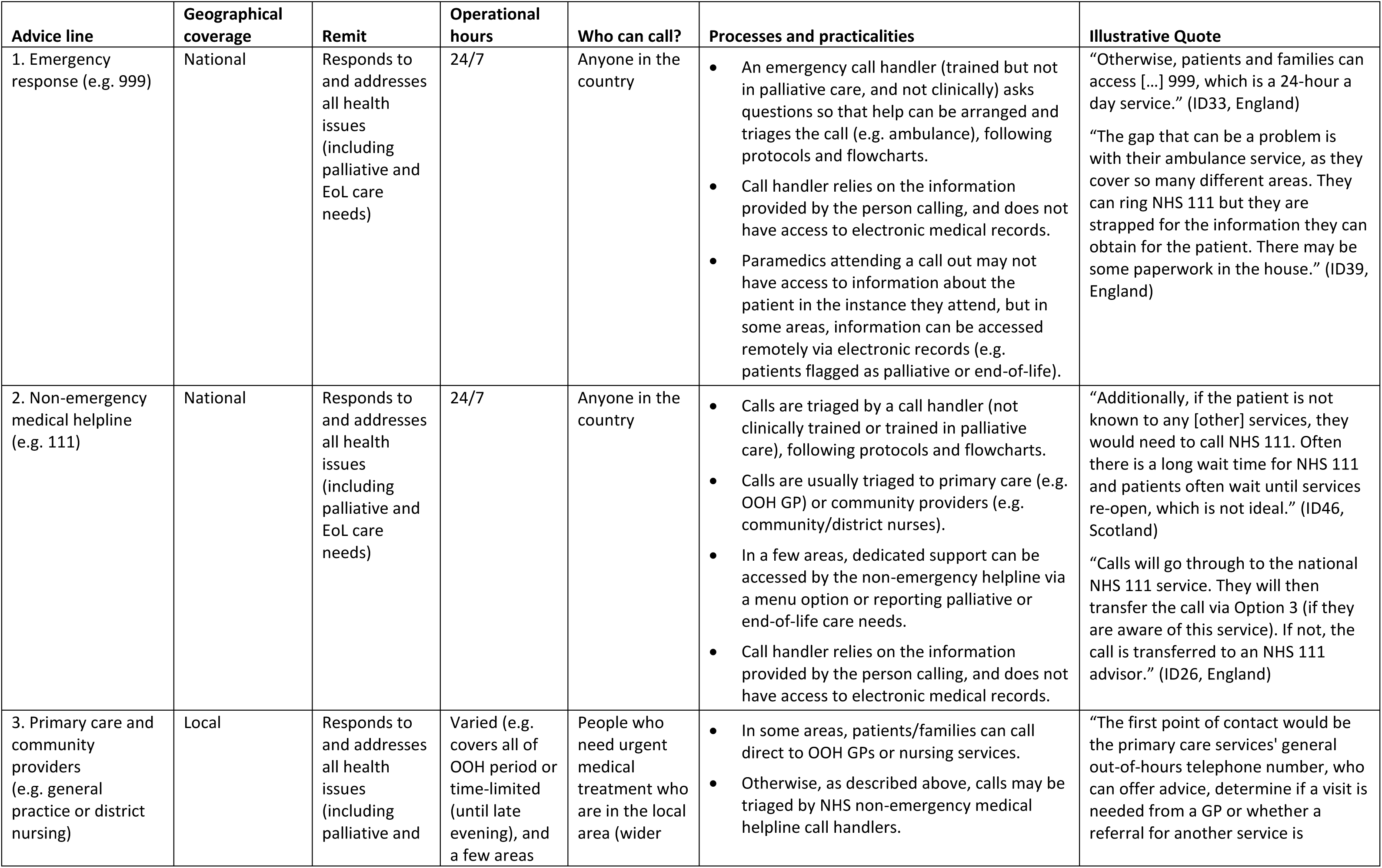
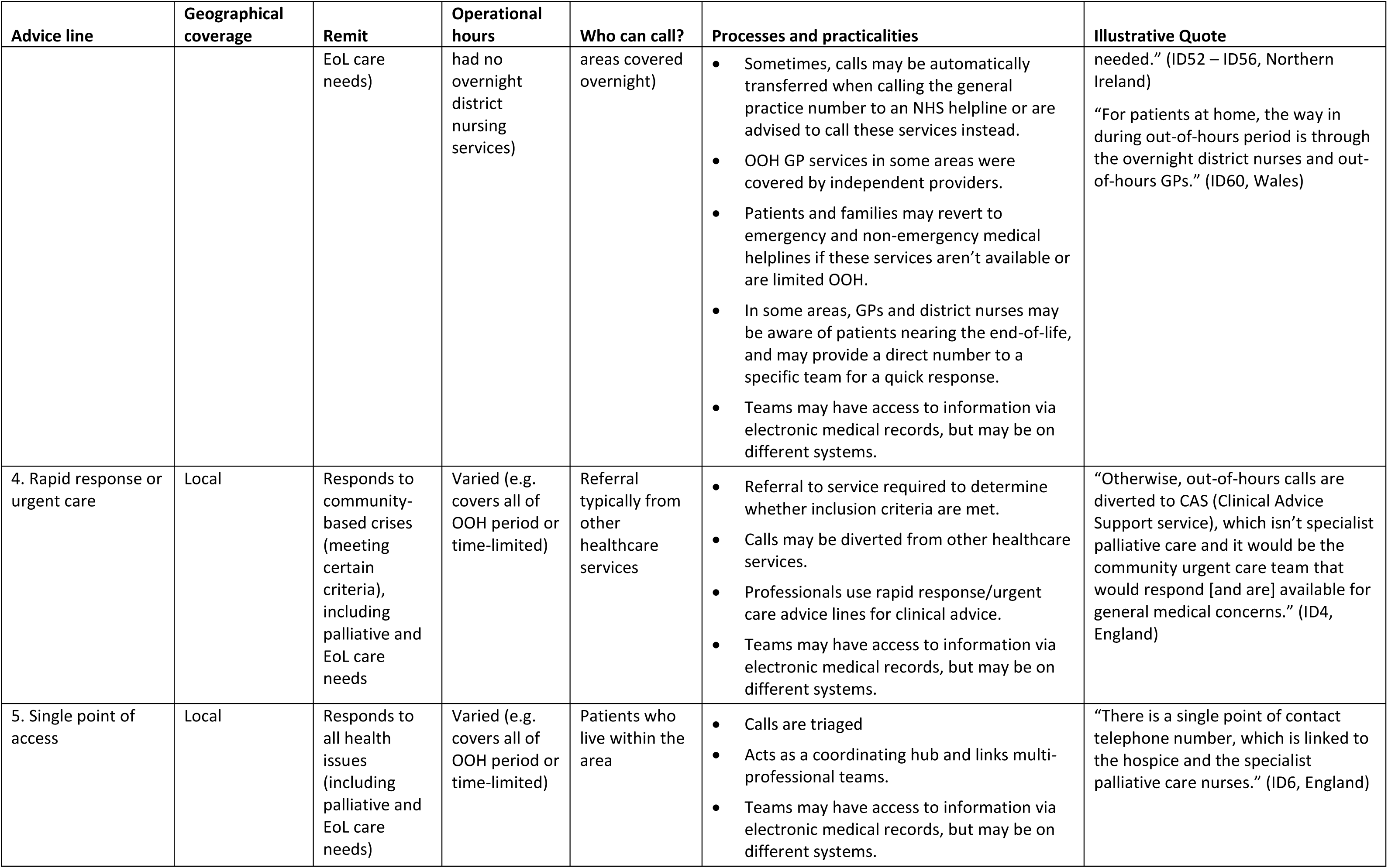
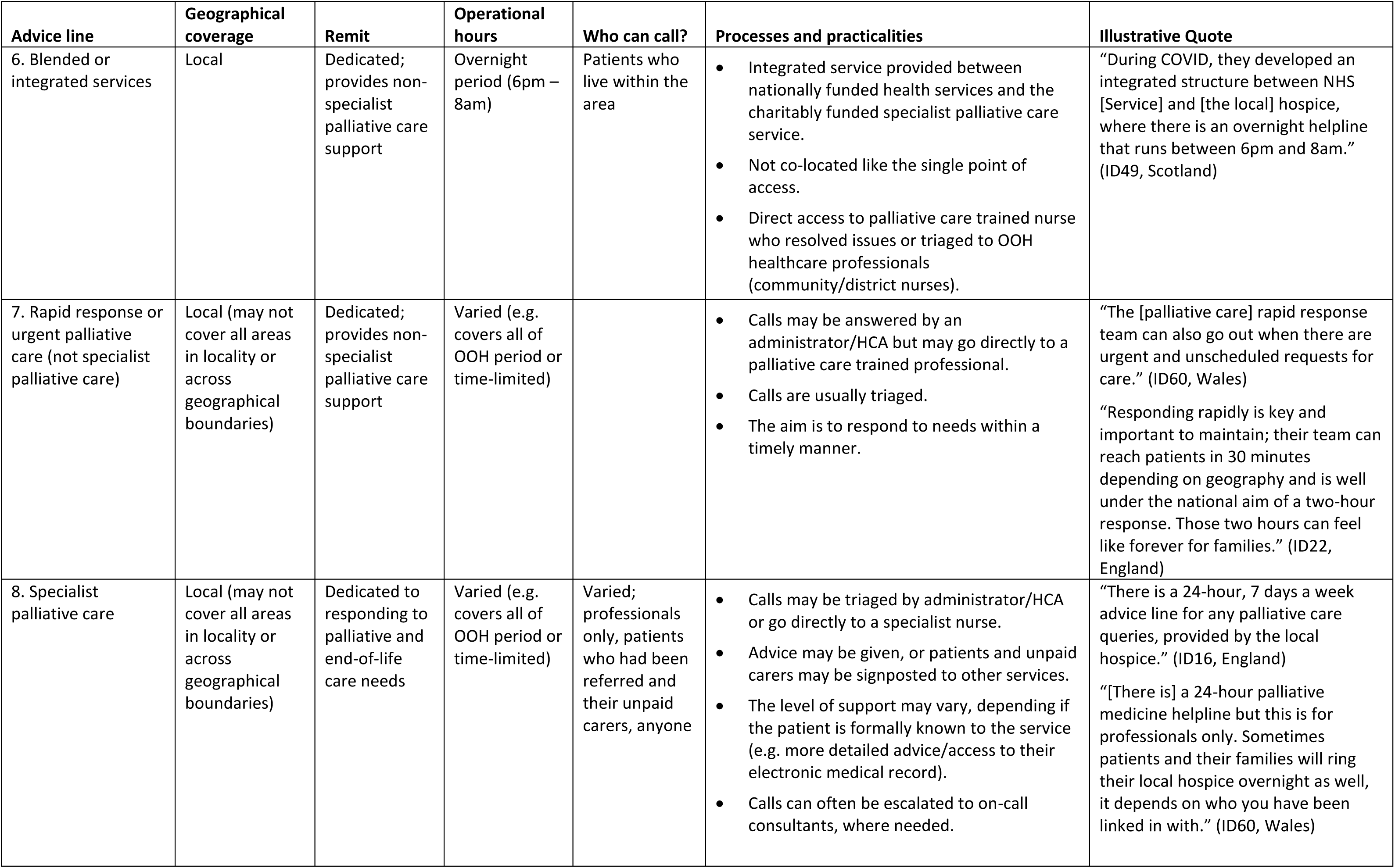
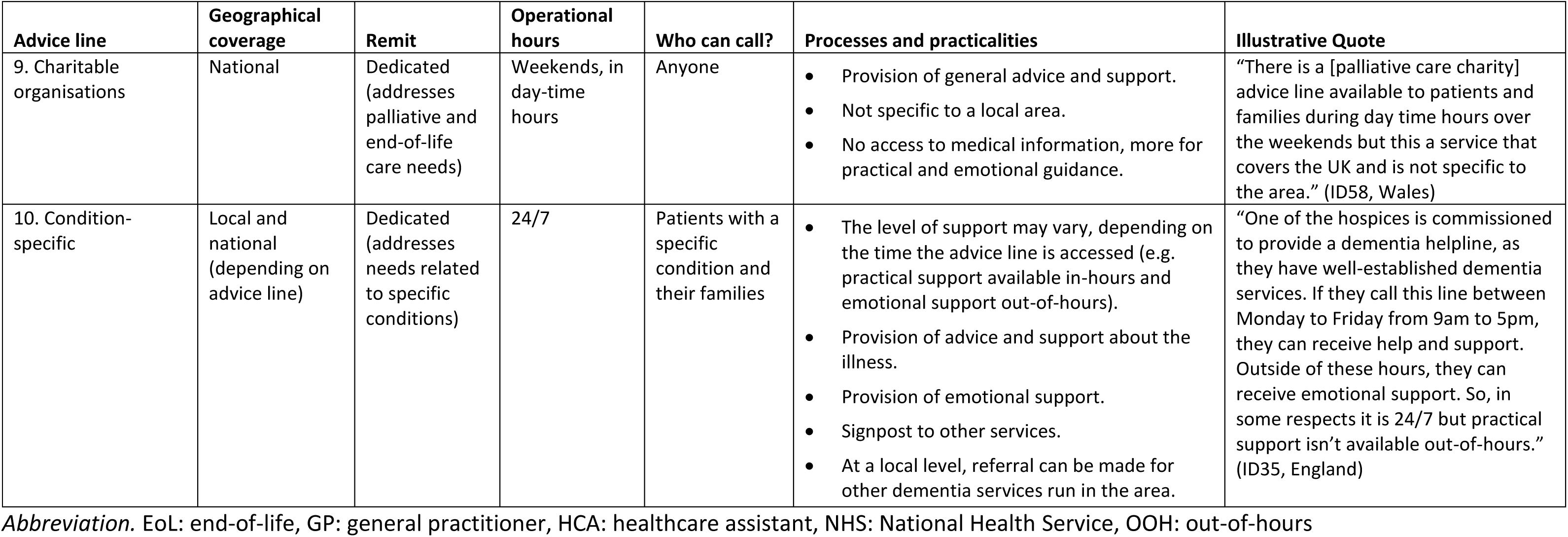
Types of advice lines, as discussed by participants.

| Advice line | Geographical coverage | Remit | Operational hours | Who can call? | Processes and practicalities | Illustrative Quote |
| --- | --- | --- | --- | --- | --- | --- |
| 1. Emergency response (e.g. 999) | National | Responds to and addresses all health issues (including palliative and EoL care needs) | 24/7 | Anyone in the country | An emergency call handler (trained but not in palliative care, and not clinically) asks questions so that help can be arranged and triages the call (e.g. ambulance), following protocols and flowcharts.<br>Call handler relies on the information provided by the person calling, and does not have access to electronic medical records.<br>Paramedics attending a call out may not have access to information about the patient in the instance they attend, but in some areas, information can be accessed remotely via electronic records (e.g. patients flagged as palliative or end-of-life). | “Otherwise, patients and families can access [...] 999, which is a 24-hour a day service.” (ID33, England)<br>“The gap that can be a problem is with their ambulance service, as they cover so many different areas. They can ring NHS 111 but they are strapped for the information they can obtain for the patient. There may be some paperwork in the house.” (ID39, England) |
| 2. Non-emergency medical helpline (e.g. 111) | National | Responds to and addresses all health issues (including palliative and EoL care needs) | 24/7 | Anyone in the country | Calls are triaged by a call handler (not clinically trained or trained in palliative care), following protocols and flowcharts.<br>Calls are usually triaged to primary care (e.g. OOH GP) or community providers (e.g. community/district nurses).<br>In a few areas, dedicated support can be accessed by the non-emergency helpline via a menu option or reporting palliative or end-of-life care needs.<br>Call handler relies on the information provided by the person calling, and does not have access to electronic medical records. | “Additionally, if the patient is not known to any [other] services, they would need to call NHS 111. Often there is a long wait time for NHS 111 and patients often wait until services re-open, which is not ideal.” (ID46, Scotland)<br>“Calls will go through to the national NHS 111 service. They will then transfer the call via Option 3 (if they are aware of this service). If not, the call is transferred to an NHS 111 advisor.” (ID26, England) |
| 3. Primary care and community providers (e.g. general practice or district nursing) | Local | Responds to and addresses all health issues (including palliative and | Varied (e.g. covers all of OOH period or time-limited (until late evening), and a few areas | People who need urgent medical treatment who are in the local area (wider | In some areas, patients/families can call direct to OOH GPs or nursing services.<br>Otherwise, as described above, calls may be triaged by NHS non-emergency medical helpline call handlers. | “The first point of contact would be the primary care services' general out-of-hours telephone number, who can offer advice, determine if a visit is needed from a GP or whether a referral for another service is |
|  |  | EoL care needs) | had no overnight district nursing services) | areas covered overnight) | • Sometimes, calls may be automatically transferred when calling the general practice number to an NHS helpline or are advised to call these services instead.<br>• OOH GP services in some areas were covered by independent providers.<br>• Patients and families may revert to emergency and non-emergency medical helplines if these services aren't available or are limited OOH.<br>• In some areas, GPs and district nurses may be aware of patients nearing the end-of-life, and may provide a direct number to a specific team for a quick response.<br>• Teams may have access to information via electronic medical records, but may be on different systems. | needed.” (ID52 – ID56, Northern Ireland)<br>“For patients at home, the way in during out-of-hours period is through the overnight district nurses and out-of-hours GPs.” (ID60, Wales) |
| 4. Rapid response or urgent care | Local | Responds to community-based crises (meeting certain criteria), including palliative and EoL care needs | Varied (e.g. covers all of OOH period or time-limited) | Referral typically from other healthcare services | • Referral to service required to determine whether inclusion criteria are met.<br>• Calls may be diverted from other healthcare services.<br>• Professionals use rapid response/urgent care advice lines for clinical advice.<br>• Teams may have access to information via electronic medical records, but may be on different systems. | “Otherwise, out-of-hours calls are diverted to CAS (Clinical Advice Support service), which isn't specialist palliative care and it would be the community urgent care team that would respond [and are] available for general medical concerns.” (ID4, England) |
| 5. Single point of access | Local | Responds to all health issues (including palliative and EoL care needs) | Varied (e.g. covers all of OOH period or time-limited) | Patients who live within the area | • Calls are triaged<br>• Acts as a coordinating hub and links multi-professional teams.<br>• Teams may have access to information via electronic medical records, but may be on different systems. | “There is a single point of contact telephone number, which is linked to the hospice and the specialist palliative care nurses.” (ID6, England) |
| 6. Blended or integrated services | Local | Dedicated; provides non-specialist palliative care support | Overnight period (6pm – 8am) | Patients who live within the area | Integrated service provided between nationally funded health services and the charitably funded specialist palliative care service.<br>Not co-located like the single point of access.<br>Direct access to palliative care trained nurse who resolved issues or triaged to OOH healthcare professionals (community/district nurses). | “During COVID, they developed an integrated structure between NHS [Service] and [the local] hospice, where there is an overnight helpline that runs between 6pm and 8am.” (ID49, Scotland) |
| 7. Rapid response or urgent palliative care (not specialist palliative care) | Local (may not cover all areas in locality or across geographical boundaries) | Dedicated; provides non-specialist palliative care support | Varied (e.g. covers all of OOH period or time-limited) |  | Calls may be answered by an administrator/HCA but may go directly to a palliative care trained professional.<br>Calls are usually triaged.<br>The aim is to respond to needs within a timely manner. | “The [palliative care] rapid response team can also go out when there are urgent and unscheduled requests for care.” (ID60, Wales)<br>“Responding rapidly is key and important to maintain; their team can reach patients in 30 minutes depending on geography and is well under the national aim of a two-hour response. Those two hours can feel like forever for families.” (ID22, England) |
| 8. Specialist palliative care | Local (may not cover all areas in locality or across geographical boundaries) | Dedicated to responding to palliative and end-of-life care needs | Varied (e.g. covers all of OOH period or time-limited) | Varied; professionals only, patients who had been referred and their unpaid carers, anyone | Calls may be triaged by administrator/HCA or go directly to a specialist nurse.<br>Advice may be given, or patients and unpaid carers may be signposted to other services.<br>The level of support may vary, depending if the patient is formally known to the service (e.g. more detailed advice/access to their electronic medical record).<br>Calls can often be escalated to on-call consultants, where needed. | “There is a 24-hour, 7 days a week advice line for any palliative care queries, provided by the local hospice.” (ID16, England)<br>“[There is] a 24-hour palliative medicine helpline but this is for professionals only. Sometimes patients and their families will ring their local hospice overnight as well, it depends on who you have been linked in with.” (ID60, Wales) |
| 9. Charitable organisations | National | Dedicated (addresses palliative and end-of-life care needs) | Weekends, in day-time hours | Anyone | • Provision of general advice and support.<br>• Not specific to a local area.<br>• No access to medical information, more for practical and emotional guidance. | "There is a [palliative care charity] advice line available to patients and families during day time hours over the weekends but this a service that covers the UK and is not specific to the area." (ID58, Wales) |
| 10. Condition-specific | Local and national (depending on advice line) | Dedicated (addresses needs related to specific conditions) | 24/7 | Patients with a specific condition and their families | • The level of support may vary, depending on the time the advice line is accessed (e.g. practical support available in-hours and emotional support out-of-hours).<br>• Provision of advice and support about the illness.<br>• Provision of emotional support.<br>• Signpost to other services.<br>• At a local level, referral can be made for other dementia services run in the area. | "One of the hospices is commissioned to provide a dementia helpline, as they have well-established dementia services. If they call this line between Monday to Friday from 9am to 5pm, they can receive help and support. Outside of these hours, they can receive emotional support. So, in some respects it is 24/7 but practical support isn't available out-of-hours." (ID35, England) |

*Abbreviation.* EoL: end-of-life, GP: general practitioner, HCA: healthcare assistant, NHS: National Health Service, OOH: out-of-hours

Availability of dedicated advice lines varied. A dedicated advice line was sometimes considered an impractical approach for managing out-of-hours care in sparsely populated regions, and statutory NHS out-of-hours services were considered able to address palliative and end-of-life care needs.

> *“They do not have one telephone line dedicated to palliative and end of life care needs/advice across the whole of North Wales for patients and their families, as it would not be practical. Patients will phone their key workers when they need support and there is always someone to call. For patients at home, the way in during out-of-hours period is through the overnight district nurses and out-of-hours GP.” ID60, Wales*

The availability of different numbers to call for out-of-hours services sometimes led to confusion about **who** patients and carers could call for help.

> *“[Patients and carers] can have a clutch of numbers, and in a crisis situation, it’s challenging to know who to ring – all of them, none of them or 999. It would depend on the family caregiver and how they cope with the situations they are managing.” ID1, England*

There was also local variation in numbers available to call between in- and out-of-hours; sometimes patients and carers needed to call a different number/select from a menu of options based on their area.

> *“[One area] are served by community nurses from [the local hospice]. These clinical nurse specialists are on-call and this team have a telephone line for their patients that can be accessed out-of-hours. For patients in [another area], they have a much smaller team covering the area, who are available from 9am until 5pm between Monday and Sunday. From 5pm until 9am, they have an arrangement that the [hospice] nurses cover their area. So, patients from [one area] will have access to the same one number, whilst patients from [the other] will have two numbers (one for in-hours Monday to Sunday, and out-of-hours).” ID42, England*

Dedicated advice lines were perceived by professionals as likely to lead to more timely care for patients with advanced illness, with generic lines sometimes leading to long waits, and taking time and effort to navigate.

> *“For patients at home, the way in during out-of-hours period is through the overnight district nurses and out-of-hours GPs. […] [The region aims for] response times [that] are within the hour but there may be some delays depending on staffing and clinical demands at that time.” ID60, Wales*

#### Times advice lines are available

Operating hours of advice lines varied. Numerous advice lines were available 24/7. These included emergency services, non-emergency medical helplines, some primary care and community nursing providers, and some dedicated advice lines. Variation in when services were available created complexity and inconsistency, and sometimes left gaps.

> *“In [the area], there is a blended service [i.e. advice line between NHS services and the hospice] with out-of-hours. This service starts at 7pm, so there is a gap between 5pm and 7pm. In [the city], the out-of-hours nursing is from 5pm until 7am. In all areas, there is a gap in services between 7am and 8am.” ID48, Scotland*

### Accessibility, awareness and promotion of advice lines

#### Accessibility

Access to dedicated advice lines was often influenced by whether a person was known to specialist palliative care services or not, and out-of-hours referral/registration were only possible in some areas.

> *“If you aren’t known, then you do get the raw deal on everything, as you just don’t have that support available. They would then just have to go via NHS 111, which would likely result in an ambulance attending and being taken to hospital when that was not necessarily what was wanted or needed.” ID35, England*

#### Awareness and promotion of advice lines

Most areas offering dedicated advice lines wanted all relevant patients and carers to know what support is available and how to access it, but recognised that this was not always achieved.

> *“There is an issue of people knowing and recognising what is available to them at home in terms of care and support. There is an assumption that someone else might inform the patient about what is available out of hours, and it is important to empower them and review their situation.” ID41, England*

Some dedicated advice lines were extensively advertised. In other areas, it was acknowledged that patients and carers would often be unaware of dedicated advice lines available, and would instead use other more general advice lines (e.g. NHS 111/999).

Nevertheless, there was an assumption that patients and carers could locate the number.

> *“The advice line would be patchy in the sense that not everyone would be aware of it. It is not necessarily advertised. Unless you were looking for it, you would not find it*.

#### This is potentially the bigger issue as there are advice lines available but people don’t always know about it.” ID11, England

Different approaches for promotion were used, including: word of mouth, websites, leaflets, care packs/folders, and local communications (for professionals to share with patients and carers).

### Practicalities of advice lines

#### Skills/expertise of call responders

The skills and expertise of call responders varied and usually related to the advice line model, with more general lines being less likely to have responders either clinically trained or trained in communicating about palliative and end-of-life needs.

On one end of the continuum were NHS call handlers, who were not clinically trained (following checklists directed towards generic health concerns) and unlikely to have received training related to palliative and end-of-life care. In contrast, dedicated advice lines were predominantly answered by nurse specialists (with different levels of seniority), who were clinically trained in palliative care. Training initiatives and resources were considered essential to support call responders in strengthening skills, competencies, and comfort, particularly for those without clinical expertise.

> *“They are currently trialling a healthcare support worker taking the first call because they find that they are better able to signpost people and solving their problems straight away. Particularly with some calls where specialist intervention is not required, where it may just be that the person has run out of continence pads or [is] unable to reach their GP. Healthcare support workers are great at getting people through to the right place.” ID7, England*

#### Processes and practical aspects

Patients and carers would call for a number of reasons, including symptom management/medications, and nursing and urgent care needs, especially when health is deteriorating. Carers were the main callers to advice lines. It was emphasised that those calling did not always need practical advice, but often reassurance and psychological support alone.

> *“If you have a well-fielded call from a family in distress, you can ensure the patient is where they need to be (such as home). A phone call can be enough to allay their fears. Out-of-hours or telephone support has an important part to play. Their team has avoided an admission by talking [about] what is happening with the family.” ID38, England*

A range of processes were described, including signposting, triaging, advice, medication management/prescribing, home visits (if needed) and follow-up. For dedicated advice lines, signposting to other services was common, especially when patients were not known to the service provider. This could lead to delays in obtaining help, but could also mean care was coordinated on the patient’s behalf.

#### A lot of the calls might be signposted elsewhere, such as GPs, hospitals and Macmillan nurses. The hospice will contact other professionals on the patient’s behalf if needed. ID16, England

Triaging was a common process across advice lines (particularly NHS 111 and some dedicated advice lines), where calls would be fielded by a telephone responder and then forwarded to an appropriate professional. Participants reflected that this sometimes delayed access to support; avoiding elongated processes was seen as essential.

> *“Out-of-hours GP’s used to have a direct number that patients could ring, including palliative care patients. They are in the process of moving over to NHS 111, which now means that calls are triaged by NHS 111 and then referred on to the appropriate service (e.g. district nursing, out-of-hours GP). ID58, Wales*

Some generic advice lines had processes which enable identification/prioritisation of patients with advanced illness, but again, this occurred only if previously known to palliative care services.

> *“If a patient calls 111 and they have a special note on their [clinical record] indicating they are palliative or end of life, they will go straight through to a clinician (instead of a call handler).” ID30, England*

Patients and carers were reported to receive more timely advice when call handlers with clinical expertise were directly accessible, which was primarily through dedicated advice lines. With dedicated advice lines, staged advice could also be provided (from nurse specialists to consultants) when calls need to be escalated. In some areas, patients and carers would seek advice by contacting their local hospice, despite these hospices not being commissioned to provide a dedicated advice line.

> *“The local hospice can be contacted for advice between 8am and 8pm, but there is less capacity for visiting and it is not a dedicated service as it is in [neighbouring area]. There is no specific helpline but patients [sometimes] call the general number of their local hospice. Otherwise, patients and families can access to 111 or 999, which is a 24-hour a day service.” ID31, England*

Where services were limited out-of-hours, there was a concern about providing advice without clinical backup if a visit is required.

> *“It was quite problematic for part of the area, as they don’t have a visiting GP service out-of-hours, and there was concern about how palliative care advice could be given, as you want to ensure there is clinical backup if a review is needed.” ID42, England*

### Integration and continuity of care

#### Collaborative working

Joined up working was seen as essential to delivering good care. Advice lines that functioned within a single point of access where multidisciplinary professionals (e.g. non-medical prescribed, specialists, and physiotherapists) were co-located were perceived as promoting joined up working between healthcare providers in the area, acted as a centralised access point, and helped guide the care delivered to patients and carers.

> *“An integrated service means that there is consistency in the advice, documentation, and guidance for palliative care and specialist palliative care across the area. So that when people ring for advice they will always get the same advice and that all the guidance is exactly the same and it only changes when it needs to and where ever people come across a patient the documentation will be the same.” ID13, England*

#### Electronic records and sharing of clinical information

The access to and sharing of a patient’s clinical information was considered essential to provide safe, timely and informed care out-of-hours.

> *“Challenges exist with the infrastructure and how the systems communicate with each other, which can delay the sharing of information.” ID57, Wales*

One particular challenge was the use of multiple electronic platforms and whether ‘read’ and/or ‘write’ permissions were given to teams working in the community, which could be particularly challenging across geographical boundaries. This meant a number of call responders would be reliant on the information provided by the patient or carer.

> *“Particularly with cross-boundary patients there can be difficulties in accessing information with a neighbouring Health Board, for example, district nursing falls under one Health Board’s remit and GP service within another.” ID57, Wales*

In some instances, dedicated platforms (electronic palliative care coordination systems) were developed to share essential information between different professions. However, these were often used variably.

> *“An electronic palliative care coordination system is currently being embedded into SystmOne and is being widely implemented so that it is freely available for everybody but there is variable use […] creating and accessing records. It is still in the implementation phase, so it is not possible to say that all providers have access to information. The hospice can register a patient on SystmOne if they get an enquiry, and setup a share but it depends on getting the necessary permissions in the other direction as well, which is challenging out-of-hours.” ID1, England*

### Service structure and commissioning

#### Local strategy and priorities

A few areas conducted mapping exercises to enable understanding of the level of support available, and how services are structured and used. However, funding to conduct such mapping activities was not always available.

> *“The ICS put a bid together to try and secure additional funding to do a mapping of what support is available via telephone, identify gaps and try and level up but their bid was unsuccessful.” ID1 and ID2, England*

Areas acknowledged the challenges with identifying patients with advanced illness to enable access to appropriate services and support.

> *“One of the avenues they are exploring for [area name] is having a single point of access for all patients, because historically the area has not been good identifying the most appropriate patients.” ID35, England*

In some areas, the development and organisation of dedicated advice lines was reported to be influenced by how palliative care was prioritised within local strategy.

> *“Over the last year and a half, they have developed their 5-year strategy plan and a dedicated out-of-hours telephone advice line is one of the key elements. [The area is] in the early scoping stages of identifying what the advice and coordination hub would look like.” ID38, England*

A few areas described the involvement of patients and carers, alongside other stakeholders, in the development of local palliative care provision, but these were the exception.

> *“There is also a clinical engagement group, which brings together the professionals involved in palliative care provision. There is also a user and carer subgroup, which brings together people who are at the end-of-life and those who care for people at the end-of-life. They have the opportunity to inform the work of the regional programme board.”. ID52 – ID56, Northern Ireland*

#### Commissioning and sustainability

Participants described the importance of access to out-of-hours palliative care, and were aware of NICE guidance on this. They recognised the need for appropriate resourcing of these services but acknowledged challenges with funding.

> *“Commissioners need to work with providers. It is very complicated in palliative and end-of-life care because much of the funding is donated via the charitable sector. It is important for commissioners to work to support both the hospice and the statutory providers to provide a NICE-compliant service.” ID41, England*

Dedicated advice lines were often provided by hospices, but were sometimes hosted by the specialist palliative care team in the community or hospital. The way advice lines were structured was considered a priority for commissioners in some areas, and sometimes lack of integration was recognised.

> *“There is a call centre that runs as part of the district nursing out-of-hours service, and [the specialist palliative care] have been challenged as to why they have a separate telephone line […]. But when this has been evaluated, it was noticed as people on that line do not have specialist knowledge, and tasks will often get passed back to [the specialist palliative care number] anyway. It then just goes a longer route round, […] their experience is that patients then receive a worse service because of this.” ID7, England*

Equitable access to dedicated advice lines was considered essential, as services that are responsive to palliative and end-of-life care needs were seen to be beneficial to other local services (e.g. community nursing services) and in improving patient and carer experiences.

> *“Equitable access to these services is so important, wherever you are. The challenges are that the community teams are so stretched, that services tend to take on additional work to support those strained services and it can become difficult to unload that work once it has been taken on. Responding rapidly is key and important to maintain; their team can reach patients in 30 minutes depending on geography and is well under the national aim of a two-hour response.” ID22, England*

Building a case for sustaining advice lines was crucial, and usually achieved via auditing/evaluation. The volume and timing of calls were considered as one possible criterion to measure service demand, and assess whether resources were used effectively. However, these varied greatly (including different of patterns across in- and out-of-hours).

> *“They are putting together a business case to get the phone line commissioned at the end of the pilot. The pilot has been well received so far. Prior to the pilot line, patients would call the single point of access line for community palliative and end-of-life services (accessed via NHS 111) but this is not available overnight.” ID3, England*

A reduction in hospital admissions were noted with dedicated advice lines. Although, challenges in capturing this were acknowledged.

### Developing a practical framework and PPI reflections

The data collected from the interviews was used to create a practical framework that presents key considerations when developing/maintaining advice lines for community-dwelling patients with advanced illness and carers. Reflections from the PPI workshop, accompanying this work, largely mirrored the discussion from the interviews with professionals, but also provided new insights. Essential and novel considerations were incorporated into the practical framework (see Figure 1 and Supplementary File 3).

**Figure 1.**
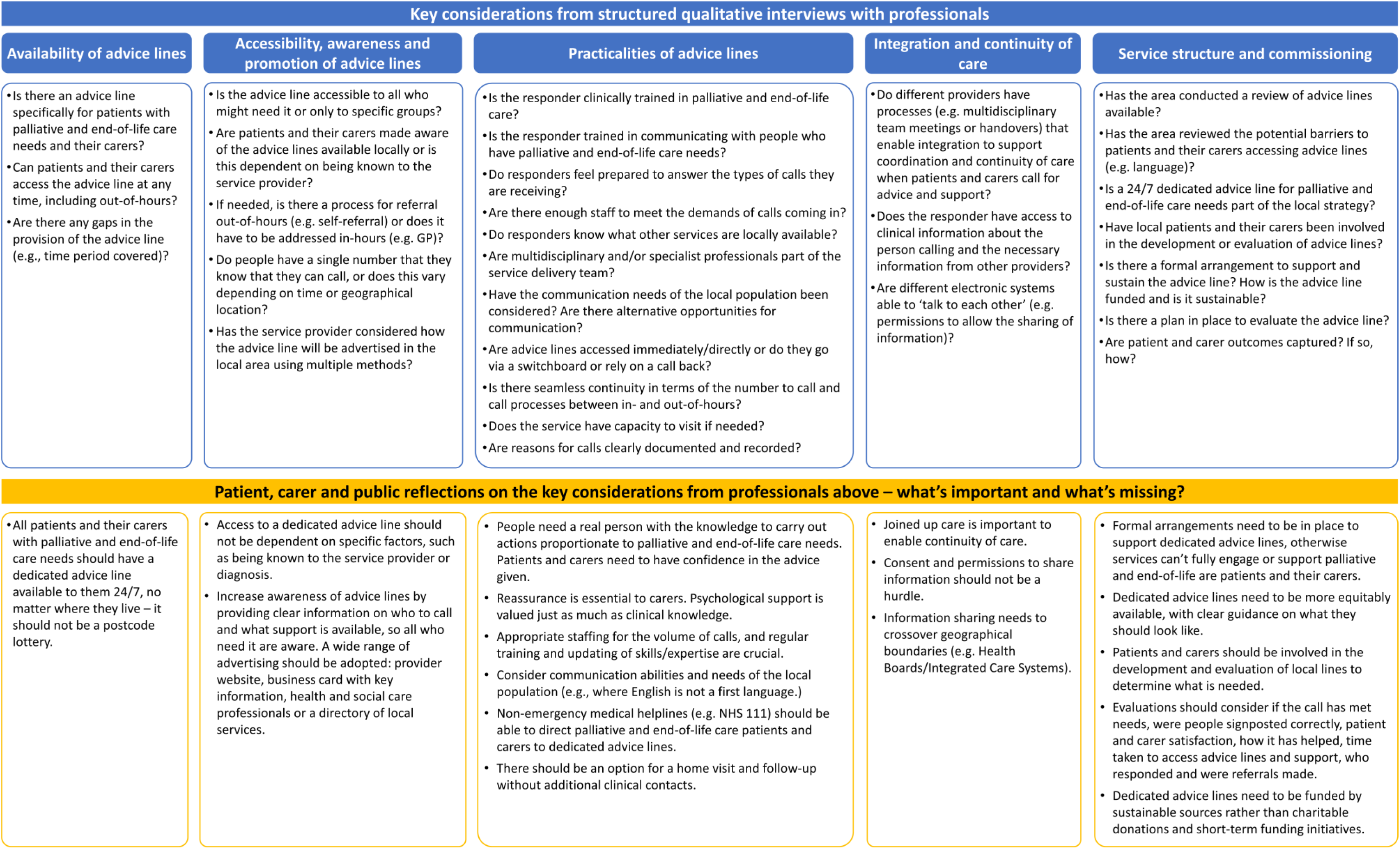
Practical framework for guiding advice lines for patients living in the community with advanced illness and their carers

## Discussion

Our findings provide, for the first time, an in-depth understanding of advice lines available out-of-hours for community-dwelling patients with advanced illness and carers at a national level. This shows the diverse models in UK provision, the variable levels of integration with other ‘out-of-hours’ services, and gaps in support. Generic advice lines are challenging for patients with advanced illness and carers to navigate. Yet there is frequent default to these as they are widely available, more accessible, and known.

Key requirements to facilitate good practice mirror previous research, including: round-the-clock availability; a single telephone number; adequate resourcing; competent professionals to answer calls; a compassionate and practical response; clear lines of responsibility where issues can be escalated; ongoing staff training and service review; access to medical records; a standardised form to collect call-related information; and advertising to raise community awareness.^32^ Our data also raises issues around identification of patients with advanced illness and referral processes, especially when self-referral is not available out-of-hours when calling dedicated advice lines. Multi-modal approaches to advertising are needed to promote services available out-of-hours. Better understanding of where palliative care is offered may improve access to this care,^33, 34^ and using novel strategies to address the multiple barriers with access and identification would be useful (e.g. reframing from a health services challenge to a community one).^35, 36^

Difficulties with accessing care in-hours has been associated with increased use of out-of-hours services.^37^ Urgent and unplanned emergency department attendance is frequent for people in the last year of life.^2, 19^ The rising demand for unscheduled care is a major burden on healthcare systems,^19^ and high-quality ‘out-of-hours’ advice lines are an important way to mitigate this.^38^ Ensuring full integration of palliative care into statutory healthcare systems, rather than parallel delivery, will enable consistency of care across settings.^39, 40^ However, investment in workforce and multidisciplinary collaboration is needed to make this happen,^41^ including training out-of-hours health and care professionals, improving sharing of clinical information, and how advice lines are evaluated (i.e. choosing meaningful patient and carer outcomes rather than focusing on service utilisation).^42–44^ Our interviews indicated that some initiatives were short-lived or developed in response to the pandemic. Palliative care services have demonstrated considerable flexibility and ‘frugal’ innovation in the context of the pandemic,^45^ but the longevity of these initiatives are important to consider.

### Strengths and limitations

Evidence on advice lines is limited, and often orientated around evaluation of single services, including in other countries.^46^ There remains a lack of robust evaluation of these types of service.^14^ One main strength of this study was providing a national picture, beyond descriptive reports of individual service use/structure. It highlights the variation and gaps in provision, as well as provides a clear structure and all advice line components. This includes how advice lines are situated in healthcare systems/services provided. Although these results may be limited in their transferability, components of this framework could have some practical applicability to other high-income countries, or potential to guide service development.

One limitation related to geographical coverage. Although we endeavoured to maximise this, it was not possible to cover all areas due to resource limitations (i.e. staffing/time). Purposive sampling was adopted to minimise bias. Another limitation was that only professionals were interviewed, as palliative care services continue to be accessed late in the course of advanced illness.^2, 47, 48^ Incorporating patients and carer priorities/preferences in the planning and development of out-of-hours palliative care services is essential to ensuring that care is patient-centred and addresses safety concerns, and has seldom been done with dedicated advice lines.^44, 49^ Therefore, we conducted a PPI workshop and integrated reflections on the evidence provided by professionals.

### Implications for clinical practice, policy and research

For the past two decades, it has been recommended that help should be available 24/7 to patients and their carers.^24^ Since, our understanding of advice lines has advanced on a small scale via audits/evaluations, and remains largely descriptive.^14, 32, 44, 50–54^ Our framework is directly informed by evidence and intended to facilitate more effective approaches to developing, enabling and evaluating such services. It may also help to understand where variations and inequalities exist. Whilst our results are based on UK data, the practical framework could be applied to other geographies to assess local provision. Further understanding is needed around the benefits and effectiveness of advice lines (e.g. cost-effectiveness), and which patient and carer outcomes to evaluate. Patients and carers need to be involved in the development, implementation and evaluation of advice lines. PPI reflections identified new aspects that require further consideration, including communication needs (such as language and cultural barriers).

### Conclusion

The multiple advice lines that can be accessed out-of-hours can lead to confusion with knowing who to call and when, and potential issues with patient safety. Dedicated advice lines are viewed as useful for streamlining and responding to palliative and end-of-life care needs, as long as they are implemented well. Currently, there is variation in the availability of dedicated advice lines, how they are accessed, promoted, and funded. The practical framework presented can provide a more robust approach to considering the essential aspects and components involved.

## Supplementary files

S1. Standards for Reporting Qualitative Research Checklist

S2. GRIPP2 – Short Form Checklist

S3. Online Facilitated Patient and Public Involvement Workshop Summary (Separate document)

## Declarations

The authors declare that there is no conflict of interest with respect to the research, authorship, or publication of this article.

## Authorship

SP, FEMM, KES and SB were responsible for the conceptualisation and design of the study, with critical input from all authors. SP conducted all structured interviews with professionals. Interviews were transcribed by SP, RLC, AM and PGM, and analysed by SP and AO, with all authors providing guidance on and the development of themes. SP was responsible for drafting and revising the manuscript. All authors critically reviewed and revised the manuscript, and read and approved the final manuscript. RK and AW led on the development of PPI activities in this study and how PPI reflections could be integrated alongside the data from professionals.

## Data Availability

Data available within the article or its supplementary materials.

## Acknowledgements

We would like to thank those who took part in the study for their time. We would also like to thank Hopkins Van Mil for contributing to the online workshop with patients and unpaid carers.

## Appendix 1 Topic guide

(1) Is there a dedicated advice line available out-of-hours for people with palliative and end-of-life care needs?
(2) Is the telephone responder clinically trained?
(3) Does the telephone responder have access to information (notes or records) regarding the person?
(4) Is there an information (electronic coordination) system available to different healthcare professionals in different care settings?

## Appendix 2 Map of geographical areas covered by interviews (from Pask et al, 2022)^2^

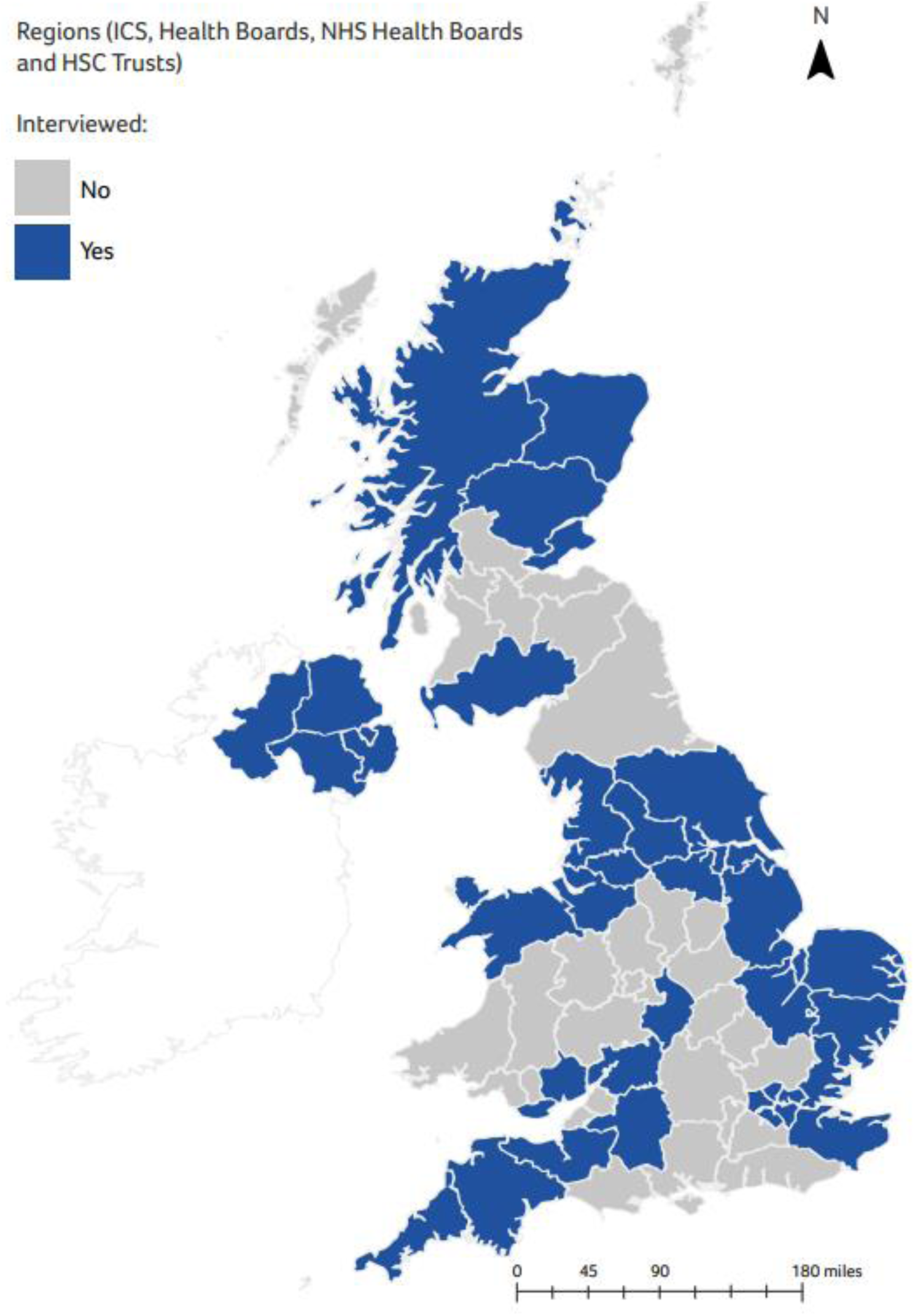

## References

1. Bone AE, Gomes B, Etkind SN, et al. What is the impact of population ageing on the future provision of end-of-life care? Population-based projections of place of death. Palliat Med 2018; 32: 329–336. 2017/10/11. DOI: 10.1177/0269216317734435.

2. Pask S, Davies JM, Mohamed A, et al. Mind the gaps: understanding and improving out-of-hours care for people with advanced illness and their informal carers (Research Report). 2022.

3. Lupati S, Stanley J, Egan R, et al. Systematic review of models of effective community specialist palliative care services for evidence of improved patient-related outcomes, equity, integration, and health service utilization. Journal of Palliative Medicine 2023. DOI: 10.1089/jpm.2022.0461.

4. Roberts B, Robertson M, Ojukwu EI, et al. Home Based Palliative Care: Known Benefits and Future Directions. Current Geriatrics Reports 2021; 10: 141–147. DOI: 10.1007/s13670-021-00372-8.

5. World Health Organisation. Why palliative care is an essential function of primary health care? 2018. https://apps.who.int/iris/handle/10665/328101.

6. Wang DH and Heidt R. Emergency Department Embedded Palliative Care Service Creates Value for Health Systems. J Palliat Med 2023; 26: 646–652. 20221111. DOI: 10.1089/jpm.2022.0245.

7. Spencer AL, Nunn AM, Miller PR, et al. The value of compassion: Healthcare savings of palliative care consults in trauma. Injury 2023; 54: 249–255. DOI: 10.1016/j.injury.2022.10.021.

8. Isenberg SR, Tanuseputro P, Spruin S, et al. Cost-effectiveness of Investment in End-of-Life Home Care to Enable Death in Community Settings. Medical Care 2020; 58.

9. Gomes B, Calanzani N, Curiale V, et al. Effectiveness and cost-effectiveness of home palliative care services for adults with advanced illness and their caregivers. Cochrane Database Syst Rev 2013; 2013: Cd007760. 20130606. DOI: 10.1002/14651858.CD007760.pub2.

10. Cassel BJ, Kerr KM, McClish DK, et al. Effect of a Home-Based Palliative Care Program on Healthcare Use and Costs. Journal of the American Geriatrics Society 2016; 64: 2288–2295. DOI: 10.1111/jgs.14354.

11. Williams H, Donaldson L, Noble S, et al. Quality improvement priorities for safer out-of-hours palliative care: Lessons from a mixed-methods analysis of a national incident-reporting database. Palliative Medicine 2019; 33: 346–356. DOI: 10.1177/0269216318817692.

12. Bliss SCAWJ. Interprofessional working in palliative care in the community: a review of the literature. Journal of Interprofessional Care 2000; 14: 281–290. DOI: 10.1080/jic.14.3.281.290.

13. Munday D, Dale J and Barnett M. Out-of-Hours Palliative Care in the UK: Perspectives from General Practice and Specialist Services. Journal of the Royal Society of Medicine 2002; 95: 28–30. DOI: 10.1177/014107680209500108.

14. Hancock S, Preston N, Jones H, et al. Telehealth in palliative care is being described but not evaluated: a systematic review. BMC Palliative Care 2019; 18: 114. DOI: 10.1186/s12904-019-0495-5.

15. Leniz J, Weil A, Higginson IJ, et al. Electronic palliative care coordination systems (EPaCCS): a systematic review. BMJ Support Palliat Care 2020; 10: 68–78. 20190508. DOI: 10.1136/bmjspcare-2018-001689.

16. Vernon E, Hughes MC and Kowalczyk M. Measuring effectiveness in community-based palliative care programs: A systematic review. Social Science & Medicine 2022; 296: 114731. DOI: 10.1016/j.socscimed.2022.114731.

17. Steeman L, Uijen M, Plat E, et al. Out-of-hours primary care in 26 European countries: an overview of organizational models. Family Practice 2020; 37: 744–750. DOI: 10.1093/fampra/cmaa064.

18. Whittaker W, Anselmi L, Kristensen SR, et al. Associations between Extending Access to Primary Care and Emergency Department Visits: A Difference-In-Differences Analysis. PLOS Medicine 2016; 13: e1002113. DOI: 10.1371/journal.pmed.1002113.

19. Mason B, Kerssens JJ, Stoddart A, et al. Unscheduled and out-of-hours care for people in their last year of life: a retrospective cohort analysis of national datasets. BMJ Open 2020; 10: e041888. 20201123. DOI: 10.1136/bmjopen-2020-041888.

20. Azhar A, Wong AN, Cerana AA, et al. Characteristics of Unscheduled and Scheduled Outpatient Palliative Care Clinic Patients at a Comprehensive Cancer Center. J Pain Symptom Manage 2018; 55: 1327–1334. 20180202. DOI: 10.1016/j.jpainsymman.2018.01.015.

21. Palliative and end-of-life care Priority Setting Partnership (PeolcPSP). Palliative and end of life care Priority Setting Partnership (PeolcPSP): Putting patients, carers and clinicians at the heart of palliative and end of life care research (Final report). London: PeolcPSP, 2015.

22. Carey ML, Zucca AC, Freund MAG, et al. Systematic review of barriers and enablers to the delivery of palliative care by primary care practitioners. Palliative Medicine 2019; 33: 1131–1145. DOI: 10.1177/0269216319865414.

23. Winkelmann J, Scarpetti G, Williams GA, et al. Policy Brief 46: How can skill-mix innovations support the implementation of integrated care for people with chronic conditions and multimorbidity? Copenhagen, Denmark: World Health Organisation, 2022.

24. National Institute for Health and Care Excellence. Guidance on cancer services: improving supportive and palliative care for adults with cancer. London, UK.2004.

25. Department of Health. End of Life Care Strategy: Promoting high quality care for all adults at the end-of-life. London: HMSO, 2008.

26. Fee A, Hasson F, Slater P, et al. Out-of-hours community palliative care: a national survey of hospice providers. Int J Palliat Nurs 2023; 29: 137–143. DOI: 10.12968/ijpn.2023.29.3.137.

27. Staniszewska S, Brett J, Simera I, et al. GRIPP2 reporting checklists: tools to improve reporting of patient and public involvement in research. Research Involvement and Engagement 2017; 3: 13. DOI: 10.1186/s40900-017-0062-2.

28. O’Brien BC, Harris IB, Beckman TJ, et al. Standards for reporting qualitative research: a synthesis of recommendations. Acad Med 2014; 89: 1245–1251. DOI: 10.1097/acm.0000000000000388.

29. Goodrich J, Tutt L, Firth AM, et al. The most important components of out-of-hours community care for patients at the end of life: A Delphi study of healthcare professionals’ and patient and family carers’ perspectives. Palliative medicine 2022; 36: 1296–1304.

30. QSR International. NVivo 14. 2023.

31. Braun V and Clarke V. Thematic Analysis: A Practical Guide. London: SAGE Publications Ltd, 2021.

32. Yardley SJ, Codling J, Roberts D, et al. Experiences of 24-hour advice line services: A framework for good practice and meeting NICE guidelines. International Journal of Palliative Nursing 2009; 15: 266–271. DOI: 10.12968/ijpn.2009.15.6.42982.

33. Gill A, Bradshaw A, Clark J, et al. 82 Public understandings of palliative care in three settings: home, hospice, hospital. BMJ Supportive & Palliative Care 2021; 11: A38–A38. DOI: 10.1136/spcare-2021-PCC.100.

34. Patel P and Lyons L. Examining the Knowledge, Awareness, and Perceptions of Palliative Care in the General Public Over Time: A Scoping Literature Review. American Journal of Hospice and Palliative Medicine® 2020; 37: 481–487. DOI: 10.1177/1049909119885899.

35. Abel J, Kellehear A, Mills J, et al. Access to palliative care reimagined. Future Healthc J 2021; 8: e699–e702. DOI: 10.7861/fhj.2021-0040.

36. Enguidanos S, Cardenas V, Wenceslao M, et al. Health Care Provider Barriers to Patient Referral to Palliative Care. American Journal of Hospice and Palliative Medicine® 2021; 38: 1112–1119. DOI: 10.1177/1049909120973200.

37. Zhou Y, Abel G, Warren F, et al. Do difficulties in accessing in-hours primary care predict higher use of out-of-hours GP services? Evidence from an English National Patient Survey. Emergency Medicine Journal 2015; 32: 373–378. DOI: 10.1136/emermed-2013-203451.

38. Etkind SN, Bone AE, Gomes B, et al. How many people will need palliative care in 2040? Past trends, future projections and implications for services. BMC Medicine 2017; 15: 102. DOI: 10.1186/s12916-017-0860-2.

39. Kamal AH, Currow DC, Ritchie CS, et al. Community-Based Palliative Care: The Natural Evolution for Palliative Care Delivery in the U.S. Journal of Pain and Symptom Management 2013; 46: 254–264. DOI: 10.1016/j.jpainsymman.2012.07.018.

40. Abel J, Kellehear A and Karapliagou A. Palliative care - the new essentials. 2018.

41. Rosa WE, Parekh de Campos A, Abedini NC, et al. Optimizing the Global Nursing Workforce to Ensure Universal Palliative Care Access and Alleviate Serious Health-Related Suffering Worldwide. Journal of Pain and Symptom Management 2022; 63: e224–e236. DOI: 10.1016/j.jpainsymman.2021.07.014.

42. Blackmore TA. What is the role of paramedics in palliative and end of life care? Palliat Med 2022; 36: 402–404. 20220217. DOI: 10.1177/02692163211073263.

43. Pocock L, Morris R, French L, et al. Underutilisation of EPaCCS (Electronic Palliative Care Coordination Systems) in end-of life-care: a cross-sectional study. BMJ Supportive & Palliative Care 2021: bmjspcare-2020-002798. DOI: 10.1136/bmjspcare-2020-002798.

44. Johansson T, Chambers RL, Curtis T, et al. A rapid review of the effectiveness of out-of-hours palliative care telephone advice lines for people living at home and their carers. Journal Article, 2023.

45. Dunleavy L, Preston N, Bajwah S, et al. ‘Necessity is the mother of invention’: Specialist palliative care service innovation and practice change in response to COVID-19. Results from a multinational survey (CovPall). Palliative Medicine 2021; 35: 814–829. DOI: 10.1177/02692163211000660.

46. Lin MH, Chen HN and Chen TJ. Changes in 24-Hour Palliative Care Telephone Advice Service after the Introduction of Discharged End-of-Life Patients’ Care Plans. Int J Environ Res Public Health 2020; 17 20200813. DOI: 10.3390/ijerph17165876.

47. McIlfatrick S, Noble H, McCorry NK, et al. Exploring public awareness and perceptions of palliative care: A qualitative study. Palliative Medicine 2014; 28: 273–280. DOI: 10.1177/0269216313502372.

48. McIlfatrick S, Slater P, Beck E, et al. Examining public knowledge, attitudes and perceptions towards palliative care: a mixed method sequential study. BMC Palliative Care 2021; 20: 44. DOI: 10.1186/s12904-021-00730-5.

49. Low C, Namasivayam P and Barnett T. Co-designing Community Out-of-hours Palliative Care Services: A systematic literature search and review. Palliative Medicine 2023; 37: 40–60. DOI: 10.1177/02692163221132089.

50. Elliker M and Barnes L. Setting up an out-of-hours palliative care helpline. Nursing Times 2008; 104: 28–29.

51. Jiang Y, Gentry AL, Pusateri M, et al. A Descriptive, Retrospective Study of After-hours Calls in Hospice and Palliative Care. J Hosp Palliat Nurs 2012; 14: 343–350.

52. Mayahara M and Fogg L. Examination and Analysis of After-Hours Calls in Hospice. Am J Hosp Palliat Care 2020; 37: 324–328. DOI: 10.1177/1049909119900377.

53. Middleton-Green L, Gadoud A, Norris B, et al. ’A Friend in the Corner’: supporting people at home in the last year of life via telephone and video consultation-an evaluation. BMJ support 2019; 9: e26. DOI: 10.1136/bmjspcare-2015-001016.

54. Wye L, Lasseter G, Percival J, et al. What works in ’real life’ to facilitate home deaths and fewer hospital admissions for those at end of life?: results from a realist evaluation of new palliative care services in two English counties. BMC Palliat Care 2014; 13: 37. DOI: 10.1186/1472-684X-13-37.

