## Supplementary file 1 for "Telephone advice lines available out-of-hours to people with palliative and end-of-life care needs: a qualitative interview study with professionals and development of a practical framework to improve services"

**SRQR checklist for conducting qualitative research^1^**

| **No.** | **Topic** | **Item** | **Page no.** | **Comments** |
| --- | --- | --- | --- | --- |
|  | *Title and abstract* | | | |
| S1 | Title | Concise description of the nature of the study, identifying the study as qualitative | p. 1 |  |
| S2 | Abstract | Summary of key elements using the abstract format of intended journal | p. 2 - 3 |  |
|  | *Introduction* | | | |
| S3 | Problem formulation | Description and significance of the problem studied | p. 4 |  |
| S4 | Purpose or research question | Purpose of the study and specific objectives | p. 4 - 5 |  |
|  | *Methods* | | | |
| S5 | Qualitative approach | Qualitative approach is described | p. 5 |  |
| S6 | Research characteristics | Researcher characteristics that may influence the research | p. 6 | See ‘Data collection’ section |
| S7 | Context | Setting/site and salient contextual factors | p. 6 | See ‘Data collection’ section |
| S8 | Sampling strategy | How and why research participants were selected | p. 6 | See ‘Sampling and recruitment’ section |
| S9 | Ethical issues | Approval from an appropriate ethics review board and participant consent, or explanation for lack thereof | p. 7 | See ‘Data collection’ and ‘Ethical approval’ sections |
| S10 | Data collection methods | Types of data collected, and collection procedures (including start and stop dates), processes, and triangulation | p. 6 | See ‘Data collection’ section |
| S11 | Data collection instruments | Description of instruments used (e.g. interview guide) and devices (e.g. audio recorder) | p. 6 and Appendix 1 | See ‘Data collection’ section and Appendix 1 |
| S12 | Units of study | Number and relevant characteristics of participants | p. 7 | See ‘Results’ section |
| S13 | Data processing | Method for processing data prior to and during analysis, including transcription, data coding, and anonymisation | p. 6 | See ‘Data collection’ section |
| S14 | Data analysis | Process by which inferences and themes were identified and developed, including research involved in the analysis. | p. 6 | See ‘Data analysis’ section |
| S15 | Techniques to enhance trustworthiness | Techniques to enhance trustworthiness and credibility of analysis (e.g. member checking) | p. 6 | See ‘Data collection’ section |
|  | *Results/findings* | | | |
| S16 | Synthesis and interpretation | Main findings (e.g. interpretations, inferences, and themes); might include development of a model or theory | p. 7 – 20 | See ‘Results’ section |
| S17 | Links to empirical data | Evidence (e.g. quotes) to substantiate findings | p. 8 – 20 | See ‘Results’ section, quotes in-text |
|  | *Discussion* | | | |
| S18 | Integration with prior work, implications, transferability and contributions to the field | Short summary of main findings, explanation of how findings and conclusions connect, support, elaborate, or challenge conclusions of earlier work, discussion of scope of application/generalisability, identification of unique contributions to field | p. 22 - 24 | See ‘Discussion’ section |
| S19 | Limitations | Trustworthiness and limitations of findings | p. 23 | See ‘Strengths and limitations’ section |
|  | *Other* | | | |
| S20 | Conflicts of interest | Potential sources of influence or perceived influence on study conduct and conclusions | p. 24 | See ‘Declarations’ section |
| S21 | Funding | Sources of funding and other support; role of funders in data collection, interpretation, and reporting | p. 25 | See ‘Funding’ section |

**Reference**

1.	O'Brien BC, Harris IB, Beckman TJ, Reed DA, Cook DA. Standards for reporting qualitative research: a synthesis of recommendations. Acad Med. 2014;89(9):1245-51.
