## Supplementary file 2 for "Telephone advice lines available out-of-hours to people with palliative and end-of-life care needs: a qualitative interview study with professionals and development of a practical framework to improve services"

**GRIPP2 – Short form checklist^1^**

| **Section and topic** | **Item** | **Reported on page number** |
| --- | --- | --- |
| 1: Aim | Report the aim of PPI in the study | Main paper, p. 6 - p. 7  Supplementary file 3, p. 1 |
| 2: Methods | Provide a clear description of the methods used for PPI in the study | Supplementary file 3, p.1 |
| 3: Study results | Outcomes—Report the results of PPI in the study, including both positive and negative outcomes | Supplementary file 3, p. 1 - p. 4 |
| 4: Discussion and conclusions | Outcomes—Comment on the extent to which PPI influenced the study overall. Describe positive and negative effects | Main paper, p. 20 (and Figure 1 on p.21)  Supplementary file 3, p. 4 |
| 5: Reflections/critical perspective | Comment critically on the study, reflecting on the things that went well and those that did not, so others can learn from this experience | Supplementary file 3, p. 4 |

**Reference**

1.	Staniszewska S, Brett J, Simera I, Seers K, Mockford C, Goodlad S, et al. GRIPP2 reporting checklists: tools to improve reporting of patient and public involvement in research. Research Involvement and Engagement. 2017;3(1):13.
