## Supplementary file 3 for "Telephone advice lines available out-of-hours to people with palliative and end-of-life care needs: a qualitative interview study with professionals and development of a practical framework to improve services"

**Summary of Patient and Public Involvement (PPI) Workshop**

**June 2023**

**Purpose of workshop**

The research presented in the main body of the paper focuses on the views of professionals. Although services are technically available, patients and carers may not be aware of them or access them easily,^1-4^ and the priorities of people using services can differ to commissioning bodies and those providing the services.^5^ Therefore, our PPI group recommended that it was important to include PPI reflections on the interviews with professionals and draw on insights from the actual experiences of people living with advanced illness and carers to expand on this data collected from professionals.

**Approach**

Patients and carers were recruited to an online facilitated workshop. Thirteen palliative and end-of-life care patients and/or carers attended; including two of our core PPI group members (representatives of England and Wales), four members of the ‘Involve Hull’ PPI network and seven members from the Cicely Saunders Institute PPI Forum (based in London). The workshop was facilitated by Hopkins Van Mil and lasted 2.5 hours (including a refreshment break). The findings from the interviews with professionals were presented by the research team in two-parts and discussed in virtual breakout rooms, and followed by a group discussion to close. The discussion from this workshop was synthesised and integrated into the practical framework presented in the main body of the paper, and have been summarised below.

***Availability of advice lines***

For patients and carers, dedicated advice lines (where available) could be an invaluable lifeline in otherwise challenging situations. Dedicated advice lines are viewed as sources of advice, knowledge, support and reassurance, which allow them to feel connected. Obtaining advice and support is more challenging out-of-hours, and it can feel though there is a countdown clock/race to request support in the transition from in-hours to out-of-hours. It was considered paramount that dedicated advice lines are available 24/7 to **all** patients and carers who need it, and should not be a ‘postcode lottery’. The specific support needed by patients with advanced illness and carers is viewed as absent where dedicated advice lines are not available.

***Accessibility, awareness and promotion of advice lines***

Access to dedicated advice and support should be equitable and not depend on circumstance (such as being already known to service providers). In reality, it is difficult for people affected by advanced illness and carers to determine what was available and when. Additionally, people do not wish to be a burden and can struggle to ask for help. A limited awareness of dedicated advice lines is a key barrier to accessing help when needed. More focus is needed on how dedicated advice lines are advertised, and when not available, how help can be sought from other services. Sometimes help may be accessed from dedicated advice lines in bordering geographical areas, but only if they are aware of them. Awareness could be raised via health and social care professionals, and services as part of routine practice to ensure that everyone with or affected by advanced illness know about this telephone-based support, and not just those recognised as palliative. This should also include non-emergency medical helplines (e.g. NHS 111) and emergency helplines (e.g. 999). In addition, adopting multiple strategies to raise awareness were also considered important. This included providing all contact information necessary when someone is recognised to be palliative, business cards with necessary information for convenience, a list of responders available, and advertising on a variety of media platforms (e.g. posters in settings of care).

***Practical aspects of advice lines***

Direct and timely access to qualified professionals who are responsive to their needs was considered the best possible outcome when calling an out-of-hours telephone-based advice line. It can be a lonely experience for carers trying to deal with a crisis and under pressure to help the person you care for. There is a dual purpose to the call responder, which includes having the necessary clinical knowledge to respond proportionately to the needs of the person calling in and being able to provide emotional/psychological support. The usual route for support is via NHS 111 or 999 but these services were not considered to be geared up to manage palliative and end-of-life care needs, and often limited by triage processes (e.g. set questions). Empathy without action when needed leads to distress, for example, when the responsibility to resolve issues is placed back onto the carer when support has not or could not be provided. Delays in care (by triage processes, leaving a voicemail, being bounced to other services, or waiting for a call back) created a reduced sense of safety. Continued training and support from out-of-hours professionals in needed.

In terms of other practical responses, the option of arranging a follow-up or home visit was considered useful if required. Additionally, specialist prescribers should be available to ensure that the necessary medicines can be provided. Self-referral could be an option out-of-hours, but it would be better if they were already engaged with palliative care services. Processes that encourage early identification of palliative care needs are useful, but only achieved in some areas (e.g. regular end-of-life care meetings within primary care). One prominent narrative that came from the workshop discussion, but not within the interviews, was that of communication. Telephone-based advice lines need to consider minoritised communities, where English is not a first language or for those who are hard-of-hearing in terms of their awareness and interaction with such services, and to enable inclusion. It is important to examine the barriers that may stop palliative care from being available to the most vulnerable in society.

***Integration and continuity of care***

Integrated care was often seen from the provider perspective rather than the patient and carer point of view. Services were often fragmented and patch-worked together, despite being considered ‘integrated’. This was particularly problematic when care is received in other geographical areas, and the information gathered there is not available in their home area. There can also be issues with access to clinical information by charitable organisations, but they are providing care to NHS patients. Access to medical records was considered vital to support a clinically safe and prompt response/record actions from the call, but currently, multiple IT systems are adopted and systems do not work together. This is a significant issue, especially when information cannot be accessed easily outside of normal working hours. Carers often felt that they had to be knowledgeable about all aspects of care, and may not know or be able to remember all information. Additionally, the repetition of information by carers when clinical information is not shared across settings added to their stress.

***Service structure and commissioning***

There is a need to standardise dedicated advice lines and how they are commissioned to ensure consistency and universal availability. There needs to be guidance on what these services should look like to support this, and ensure that there is access to appropriate professionals who can help. Services need to perform to basic guidelines and require support with funding to achieve this. It was challenging for patients and carers to understand why these services were not statutorily funded. Charitable funding is diminished and should not vary by affluence, where charitable donations may be higher. It is important that all areas have the same level of services. Some issues are created by the way we structure and organise services, such as issues with crossover between geographical areas and how palliative and end-of-life care is prioritised within strategies. Ongoing and routine monitoring is essential to determine what is and is not working. Evaluations should follow-up with patients and/or their carers to determine how the advice line met their needs and who by, the time taken to access the service, actions from the call, impact on other service use, and whether it was helpful.

***Reflections***

The input from patients and carers was invaluable as patients and carers are seldom involved in the development and evaluation of such services, and allowed us to confirm and expand on findings from professionals. The use of a service to facilitate the workshop was of great benefit and meant that the research team was able to be more present in the discussion with patients and carers. Although, this does require appropriate funds to support. The online element allowed for people to join remotely and supported breakout discussions. We managed to include thirteen PPI members, but there may views and experiences missed that could further shape the framework.

**References**

1.	Pask S, Davies JM, Mohamed A, Javiera L, Chambers RL, McFarlane P, et al. Mind the gaps: understanding and improving out-of-hours care for people with advanced illness and their informal carers (Research Report). 2022.

2.	McIlfatrick S, Noble H, McCorry NK, Roulston A, Hasson F, McLaughlin D, et al. Exploring public awareness and perceptions of palliative care: A qualitative study. Palliative Medicine. 2014;28(3):273-80.

3.	McIlfatrick S, Slater P, Beck E, Bamidele O, McCloskey S, Carr K, et al. Examining public knowledge, attitudes and perceptions towards palliative care: a mixed method sequential study. BMC Palliative Care. 2021;20(1):44.

4.	Patel P, Lyons L. Examining the Knowledge, Awareness, and Perceptions of Palliative Care in the General Public Over Time: A Scoping Literature Review. American Journal of Hospice and Palliative Medicine®. 2020;37(6):481-7.

5.	Goodrich J, Tutt L, Firth AM, Evans CJ, Murtagh FEM, Harding R. The most important components of out-of-hours community care for patients at the end of life: A Delphi study of healthcare professionals’ and patient and family carers’ perspectives. Palliative medicine. 2022;36(8):1296-304.
